# Routine saliva testing for the identification of silent COVID-19 infections in healthcare workers

**DOI:** 10.1101/2020.11.27.20240044

**Authors:** Kevin Zhang, Affan Shoukat, William Crystal, Joanne M. Langley, Alison P. Galvani, Seyed M. Moghadas

## Abstract

**Objective:** Current COVID-19 guidelines recommend symptom-based screening and regular nasopharyngeal (NP) testing for healthcare personnel in high-risk settings. We sought to estimate case detection percentages with various routine NP and saliva testing frequencies.

**Design:** Simulation modelling study.

**Methods:** We constructed a sensitivity function based on the average infectiousness profile of symptomatic COVID-19 cases to determine the probability of being identified at the time of testing. This function was fitted to reported data on the percent positivity of symptomatic COVID-19 patients using NP testing. We then simulated a routine testing program with different NP and saliva testing frequencies to determine case detection percentages during the infectious period, as well as the pre-symptomatic stage.

**Results:** Routine bi-weekly NP testing, once every two weeks, identified an average of 90.7% (SD: 0.18) of cases during the infectious period and 19.7% (SD: 0.98) during the pre-symptomatic stage. With a weekly NP testing frequency, the corresponding case detection percentages were 95.9% (SD: 0.18) and 32.9% (SD: 1.23), respectively. A 5-day saliva testing schedule had a similar case detection percentage as weekly NP testing during the infectious period, but identified about 10% more cases (mean: 42.5%; SD: 1.10) during the pre-symptomatic stage.

**Conclusion:** Our findings highlight the utility of routine non-invasive saliva testing for frontline healthcare workers to protect vulnerable patient populations. A 5-day saliva testing schedule should be considered to help identify silent infections and prevent outbreaks in nursing homes and healthcare facilities.

## Introduction

The novel coronavirus disease 2019 (COVID-19) has led to a devastating global pandemic ^1^. The burden of disease has been disproportionately high in some healthcare settings and long-term care facilities, with case fatality rates exceeding 30% ^2,3^. Most COVID-19 cases among healthcare workers are the result of community exposure ^4^, posing a potential risk of transmission to immunocompromised individuals and those at higher risk of developing adverse clinical outcomes ^5–8^. Modelling analyses show that rapid case identification of infected persons is critical to interrupt transmission, especially for infectious cases without clinical symptoms ^9^.

Current case detection approaches in healthcare settings rely on symptom-based screening and nasopharyngeal (NP) testing for symptomatic or exposed healthcare personnel ^10,11^. Some jurisdictions have recommended routine bi-weekly or weekly NP testing for frontline healthcare workers in facilities at risk of severe COVID-19 outbreaks, such as nursing homes ^10,11^. The nasopharyngeal test to detect nucleic acid or antigen, however, is relatively invasive and requires trained healthcare providers for sample collection. On the other hand, saliva tests can be self-administered, and therefore are easier to implement, potentially more acceptable, and reduce the need for personal protective equipment (PPE) during sample collection ^12^. Since up to 80% of COVID-19 cases are mild or asymptomatic ^13^ and therefore might be missed by symptom based screening, testing of asymptomatic healthcare personnel could increase detection and prevent transmission during the highly infectious pre-symptomatic period ^9,14^. An easy-to-administer saliva test could be a more feasible tool to conduct higher frequency testing to curtail silent transmission. A recent modelling study suggests that at least 33% of silent infections must be identified and isolated in the pre-symptomatic or asymptomatic stage of the disease to enable outbreak control, even when all symptomatic cases are immediately isolated^9^.

Given the importance of testing in preventing onward transmission in healthcare settings, we sought to estimate case detection percentages using RT-PCR testing of NP and saliva samples, and ascertain the frequency of testing that may be required to control outbreaks.

## Methods

We simulated a routine testing program with various frequencies of NP and saliva tests over a time horizon of 150 days. In our analysis, we only included individuals who went on to develop a symptomatic course of disease. To estimate case detection percentages, we first constructed a sensitivity function *s*_*τ*_(*t*) to map the infectiousness profile of symptomatic COVID-19 cases ^14,15^ to the reported percent positivity of NP RT-PCR tests post-symptom onset ^16^. The infectiousness profile (Appendix: Figure A1) was extracted from computer code provided in previous studies that utilized maximum likelihood and optimization methods ^14,15^. The mapping was performed by fitting the sensitivity function to the publicly available percent positivity data of 209 COVID-19 patients for 26 days after the start of symptoms, including the day of symptom onset ^16^. The sensitivity function, expressed as the product of Hill and Gompertz functions, is given by

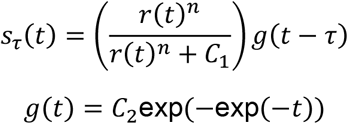

where *r*(*t*) is the average infectiousness profile over time *t, n* is the Hill coefficient, *C*_1_ is the Hill saturation constant, and *C*_2_ is the Gompertz asymptote level. The parameter *τ* indicates the start of infectiousness, which was assumed to be one day after infection within the incubation period. For each infected individual, the incubation period was sampled independently from a LogNormal distribution, with parameters 1.434 (shape) and 0.661 (rate), having a mean of 5.2 days ^17^.

We fitted the sensitivity function using a least-squares method, and obtained time-dependent NP RT-PCR sensitivities for different values of *τ*, which determined the probability of being detected at the time of an NP test. Given the timelines of infectiousness profile (Appendix: Figure A1), we considered a detection period from the start of infectiousness to 15 days post-symptom onset as clinically relevant for disease transmission. The case detection percentage was then calculated as the average probability of all individuals being identified in at least one test within their infectious period. To determine the case detection percentage with a saliva test, we used recent empirical studies for the estimates of saliva testing sensitivity in the range of 70% - 97% ^18–20^. Since viral load in saliva samples have shown to be comparable to NP samples over time ^21–23^, we applied this range to the normalized sensitivity curves of NP testing and determined the temporal sensitivity of a saliva test (Appendix: Figure A2). Normalization was done by dividing each point on the fitted NP sensitivity curve by its maximum estimated sensitivity over time. Further details of the model implementation are provided in the Appendix.

To derive the distributions for mean case detection percentages during the infectious period and the pre-symptomatic stage, we ran 500 independent Monte-Carlo simulations by introducing 100 infections on each day for each simulation. The generated distributions were then compared using the Mann Whitney U test. We conducted this analysis to ascertain the frequencies of testing needed to identify at least one-third, one-half, and two-thirds of silent infections during the pre-symptomatic stage.

### Ethics approval

This research was based on publicly available data ^14–16^ and therefore did not require ethics approval.

## Results

### Impact of routine testing on infectious case identification

Biweekly NP testing, once every two weeks, identified, on average, 90.7% (SD: 0.18) of cases during the infectious period (Figure 1A). With a weekly NP testing schedule, the case detection percentage was 95.9% (SD: 0.18) (Figure 1B). We found that 81.2% of individuals were detected by the first NP test, irrespective of the testing frequency. Biweekly saliva testing identified a mean of 78.6% (SD: 0.24) of cases during the infectious period (Figure 1C). When the frequency of saliva testing increased to a weekly schedule, the case detection percentage was 91.2% (SD: 0.24) (Figure 1D). With saliva testing, the detection percentage for the first test was 67.8%, irrespective of the testing frequency.

**Figure 1.**
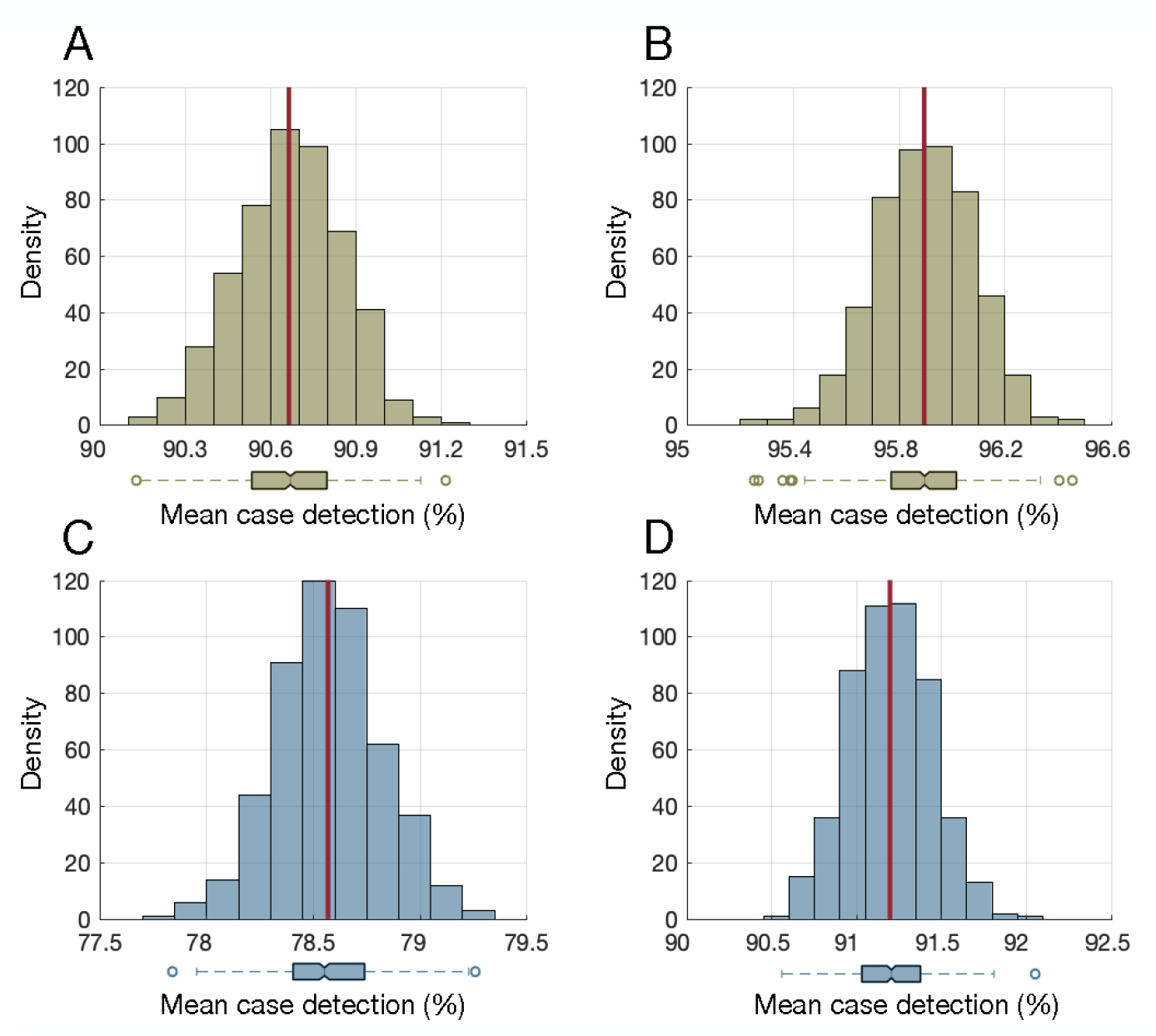
Distribution of mean case detection percentages during the infectious period using bi-weekly nasopharyngeal (A) and saliva (C) testing. Distribution of mean case detection percentages during the infectious period using weekly nasopharyngeal (B) and saliva (D) testing. The red line indicates the mean of the distribution, and the boxplot represents the interquartile range (IQR) with whiskers extending the range from minimum (25th percentile – 1.5 IQR) to maximum (75th percentile + 1.5 IQR). The density on the y-axis is the number of experiments from 500 iterations (Monte-Carlo simulations) that resulted in a mean case detection shown on the x-axis.

An 8-day saliva testing frequency was required to identify a similar percentage of infectious cases as with NP testing every two weeks, with no significant difference between the distributions of mean detection percentages in the two tests (Mann Whitney U test, p-value=0.33). A frequency of 5-day saliva testing had a similar infectious case detection percentage as compared to weekly NP testing, with no significant difference in the distributions of mean detection percentages between the two tests (Mann Whitney U test, p-value=0.16).

### Impact of routine testing on pre-symptomatic case identification

Biweekly NP testing identified an average of 19.7% (SD: 0.98) of cases during the pre-symptomatic infectious stage (Figure 2A). With a weekly NP testing schedule, the mean pre-symptomatic case detection percentage was 32.9% (SD: 1.23) (Figure 2B). For saliva testing, the mean case detection percentages during the pre-symptomatic stage were 16.4% (SD: 0.83) and 32.4% (SD: 1.10) for biweekly and weekly schedules, respectively (Figures 2C, 2D). A 5-day saliva testing schedule, while detecting a similar percentage of cases as weekly NP testing during the infectious period, identified a mean of 42.5% (SD: 1.10) of pre-symptomatic cases, which was significantly different (Mann Whitney U test, p-value<0.001) and about 10% higher than weekly NP testing.

**Figure 2.**
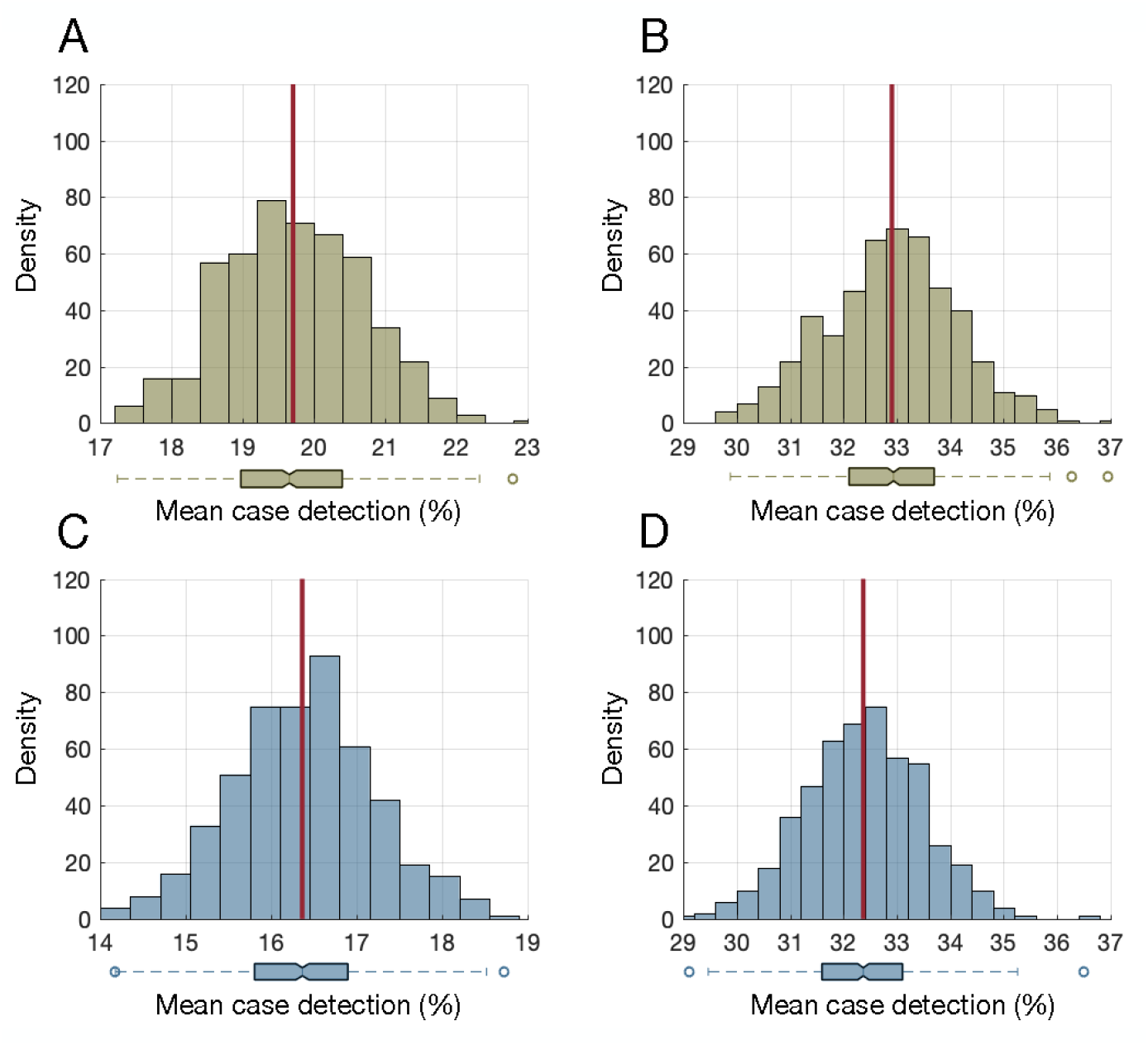
Distribution of mean case detection percentages during the pre-symptomatic stage using bi-weekly nasopharyngeal (A) and saliva (C) testing. Distribution of mean case detection percentages during the pre-symptomatic stage using weekly nasopharyngeal (B) and saliva (D) testing. The red line indicates the mean of the distribution, and the boxplot represents the interquartile range (IQR) with whiskers extending the range from minimum (25th percentile – 1.5 IQR) to maximum (75th percentile + 1.5 IQR). The density on the y-axis is the number of experiments from 500 iterations (Monte-Carlo simulations) that resulted in a mean case detection shown on the x-axis.

A routine NP testing frequency of at least once every 6 days, 4 days, and 2 days was required for the case detection percentage during the pre-symptomatic stage to exceed 33%, 50%, and 67%, respectively. We found that the same saliva testing frequencies would be required to exceed pre-symptomatic case detection percentages of 33%, 50%, and 67%, respectively.

## Discussion

Our results show that routine NP testing every two weeks or every week, as recommended by some jurisdictions for frontline healthcare workers ^10,11^, would lead to a significant percentage of undetected silent COVID-19 infections, indicating that institutional outbreaks could occur even in the presence of symptom-based screening ^9^. Recent studies suggest that a significant portion of disease transmission occurs prior to symptom onset ^14,24,25^, highlighting the importance of early detection. Given the practical considerations with NP testing, non-invasive saliva testing presents an attractive alternative for improving case detection with increased testing frequency ^26,27^. Moreover, despite a higher sensitivity, the more invasive NP test did not reduce the required frequency of testing to identify at least 33%, 50%, and 67% of cases in the pre-symptomatic stage.

Until vaccines are available, healthcare settings and long-term care facilities remain vulnerable to outbreaks that could be seeded through silent transmission by asymptomatic or pre-symptomatic healthcare personnel. Adherence to public health measures, behavioural interventions and standard and additional precautions will be essential. Routine testing is an additional intervention that could, along with early case detection of infected healthcare workers, prevent the introduction of COVID-19 to healthcare settings ^21^. NP testing, while more sensitive compared to saliva testing, is relatively invasive and requires trained personnel to sample individuals, making frequent NP testing impractical for large scale implementation. For example, a recent study suggests that a testing frequency of every 2 days with a test sensitivity above 70% would be needed to prevent outbreaks in post-secondary settings ^28^. Given the high frequency of testing required to detect a sufficient number of silent infections to prevent outbreaks, compliance rates would likely be higher with a non-invasive saliva test.

Our study was based on the assumption of infection *a priori*, and therefore, did not estimate false positive rates. Given the high specificity of NP and saliva testing estimated at 99.93% (90%CI: 99.77%-99.99%) and 99.96% (90%CI: 99.85%-100.00%), respectively ^18^, false positive rates would vary depending on the test frequency but are likely to remain under 2%. For instance, if tests are done every two weeks, with a maximum of 3 tests conducted during the infectious period, *m =* 3, the false positivity rate could reach (1 − (1 − *F* _*p*_)^*m*^)100% *=* 0.69% for an upper-bound test false positivity *Fp =* 0.0023 (given a specificity of 99.77%). However, this may still lead to a slightly higher rate of self-isolation than necessary compared to a test with perfect specificity, as current guidelines recommend that healthcare personnel be excluded from work for 14 days following a known exposure or positive test ^10,11,29^. In our analysis, we only included individuals who went on to develop a symptomatic course of disease. However, given that recent studies have shown similar viral loads for asymptomatic and symptomatic cases ^30,31^, we expect that our case detection estimates would be applicable for detecting asymptomatic individuals during the infectious period. We also did not model the effect of contact tracing which would readily identify individuals for testing based on known exposures and impose self-isolation if test results are available in a timely manner. In a real-life setting, when contact-tracing is combined with routine testing and appropriate referrals are made to a more sensitive NP test as required, the effectiveness of a routine testing strategy would be enhanced. Finally, in order to evaluate the independent impact of a routine testing strategy, we did not consider other mitigation measures.

Our findings highlight the importance and utility of routine non-invasive saliva testing for frontline healthcare workers in order to protect vulnerable patient populations. Coupled with contact tracing and infection prevention and control measures, a 5-day routine saliva testing schedule presents an attractive screening method to reduce the risk of outbreaks in healthcare settings.

## Data Availability

The computational model with parameter values and data pertaining to the simulation study are freely available at: https://github.com/affans/npt-saliva-testing.

## Contributors

Kevin Zhang, Joanne Langley, Alison Galvani, and Seyed Moghadas contributed to the conception and design of the work. Kevin Zhang, Affan Shoukat, William Crystal, and Seyed Moghadas contributed to the acquisition of data, analysis, and interpretation of results. Affan Shoukat performed the simulations. All of the authors drafted the manuscript and approved it.

## References

1. Johns Hopkins University. COVID-19 Map. Johns Hopkins Coronavirus Resource Center.

2. McMichael TM, Currie DW, Clark S, et al. Epidemiology of Covid-19 in a Long-Term Care Facility in King County, Washington. N Engl J Med. 2020;382(21):2005–2011.

3. Comas-Herrera A, Zalakaín J, Lemmon E, et al. Mortality associated with COVID-19 outbreaks in care homes: early international evidence.

4. Alberta Health Services. COVID-19 Scientific Advisory Group Rapid Response Report.

5. Wang B, Li R, Lu Z, Huang Y. Does comorbidity increase the risk of patients with COVID-19: evidence from meta-analysis. Aging (Albany NY). 2020;12(7):6049–6057.

6. Kim L, Garg S, O’Halloran A, et al. Risk Factors for Intensive Care Unit Admission and In-hospital Mortality Among Hospitalized Adults Identified through the US Coronavirus Disease 2019 (COVID-19)-Associated Hospitalization Surveillance Network (COVID-NET). Clin Infect Dis. Published online July 16, 2020.

7. Garnier-Crussard A, Forestier E, Gilbert T, Krolak-Salmon P. Novel Coronavirus (COVID-19) Epidemic: What Are the Risks for Older Patients? Journal of the American Geriatrics Society. 2020;68(5):939–940.

8. Yang J, Zheng Y, Gou X, et al. Prevalence of comorbidities and its effects in patients infected with SARS-CoV-2: a systematic review and meta-analysis. International Journal of Infectious Diseases. 2020;94:91-95.

9. Moghadas SM, Fitzpatrick MC, Sah P, et al. The implications of silent transmission for the control of COVID-19 outbreaks. PNAS. Published online July 6, 2020.

10. Centers for Disease Control and Prevention. Interim Guidance on Testing Healthcare Personnel for SARS-CoV-2.

11. Ontario Health. COVID-19 Surveillance Testing - Guidance Regarding Retirement Homes Staff and Resident Testing.

12. Public Health Ontario. The use of saliva as an alternate specimen for SARS-CoV-2 (COVID-19) PCR testing.

13. World Health Organization. Coronavirus disease 2019 (COVID-19) Situation Report - 46.

14. He X, Lau EHY, Wu P, et al. Temporal dynamics in viral shedding and transmissibility of COVID-19. Nature Medicine. 2020;26(5):672–675.

15. Ashcroft P, Huisman JS, Lehtinen S, et al. COVID-19 infectivity profile correction. Swiss Medical Weekly. 2020;150(3132).

16. Miller TE, Beltran WFG, Bard AZ, et al. Clinical sensitivity and interpretation of PCR and serological COVID-19 diagnostics for patients presenting to the hospital. The FASEB Journal. 2020;34(10):13877–13884.

17. Li Q, Guan X, Wu P, et al. Early Transmission Dynamics in Wuhan, China, of Novel Coronavirus–Infected Pneumonia. N Engl J Med. 2020;382(13):1199–1207.

18. Yokota I, Shane PY, Okada K, et al. Mass screening of asymptomatic persons for SARS-CoV-2 using saliva. Clin Infect Dis. Published online September 25, 2020.

19. Jamal AJ, Mozafarihashjin M, Coomes E, et al. Sensitivity of Nasopharyngeal Swabs and Saliva for the Detection of Severe Acute Respiratory Syndrome Coronavirus 2. Clin Infect Dis. Published online June 25, 2020.

20. Wyllie AL, Fournier J, Casanovas-Massana A, et al. Saliva or Nasopharyngeal Swab Specimens for Detection of SARS-CoV-2. N Engl J Med. 2020;383(13):1283–1286.

21. Larremore DB, Wilder B, Lester E, et al. Test Sensitivity Is Secondary to Frequency and Turnaround Time for COVID-19 Surveillance. medRxiv; 2020.

22. Iwasaki S, Fujisawa S, Nakakubo S, et al. Comparison of SARS-CoV-2 detection in nasopharyngeal swab and saliva. J Infect. 2020;81(2):e145–e147.

23. Kim SE, Lee JY, Lee A, et al. Viral Load Kinetics of SARS-CoV-2 Infection in Saliva in Korean Patients: a Prospective Multi-center Comparative Study. J Korean Med Sci. 2020;35(31):e287.

24. Ren X, Li Y, Yang X, et al. Evidence for pre-symptomatic transmission of coronavirus disease 2019 (COVID-19) in China. Influenza and Other Respiratory Viruses. Published online August 7, 2020.

25. Casey M, Griffin J, McAloon CG, et al. Pre-Symptomatic Transmission of SARS-CoV-2 Infection: A Secondary Analysis Using Published Data. medRxiv; 2020.

26. Moreno-Contreras J, Espinoza MA, Sandoval-Jaime C, et al. Saliva Sampling and Its Direct Lysis, an Excellent Option To Increase the Number of SARS-CoV-2 Diagnostic Tests in Settings with Supply Shortages. Caliendo AM, ed. J Clin Microbiol. 2020;58(10).

27. Wei S, Kohl E, Djandji A, et al. Field-Deployable, Rapid Diagnostic Testing of Saliva Samples for SARS-CoV-2. medRxiv; 2020.

28. Paltiel AD, Zheng A, Walensky RP. Assessment of SARS-CoV-2 Screening Strategies to Permit the Safe Reopening of College Campuses in the United States. JAMA Netw Open. 2020;3(7):e2016818.

29. Public Health Agency of Canada. COVID-19: How to quarantine (self-isolate) at home when you may have been exposed and have no symptoms.

30. Ra SH, Lim JS, Kim G, Kim MJ, Jung J, Kim S-H. Upper respiratory viral load in asymptomatic individuals and mildly symptomatic patients with SARS-CoV-2 infection. Thorax. Published online September 22, 2020.

31. Lee S, Kim T, Lee E, et al. Clinical Course and Molecular Viral Shedding Among Asymptomatic and Symptomatic Patients With SARS-CoV-2 Infection in a Community Treatment Center in the Republic of Korea. JAMA Intern Med. Published online August 6, 2020.

