## Supplementary File for "Routine saliva testing for the identification of silent COVID-19 infections in healthcare workers"

This appendix includes the infectiousness profile and details of the fitting of the modified Hill function, corresponding to the results reported in the main text.

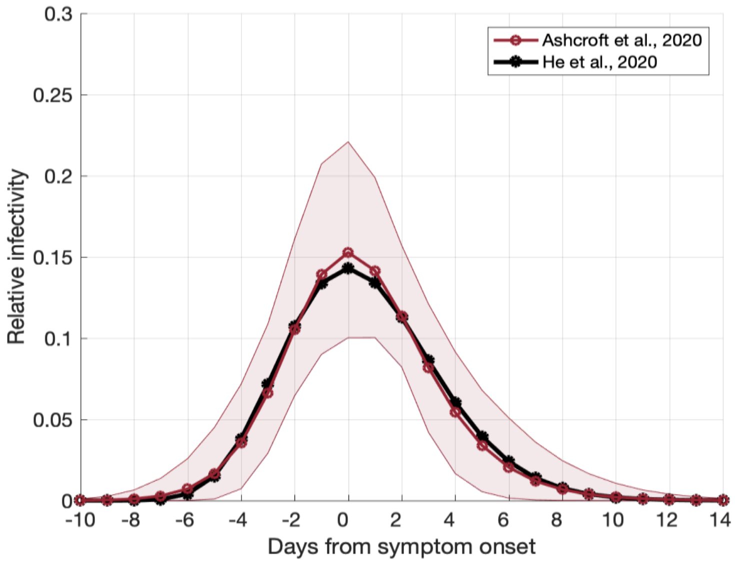


**Figure A1.** Distribution of the infectiousness profile. The distributions are generated using parameters extracted from computer code provided in previous studies that utilized maximum likelihood and optimization methods [1,2].

#### Construction of the sensitivity function

The sensitivity function $s_\tau(t)$ is expressed as the product of Hill and Gompertz functions given by

$$s_\tau(t) = \left(\frac{r(t)^n}{r(t)^n + C_1}\right) g(t-\tau)$$

$$g(t) = C_2\text{exp}(-\text{exp}(-t))$$

where $r(t)$ is the average infectiousness profile over time $t$ (Figure A1), $n$ is the Hill coefficient, $C_1$ is the Hill saturation constant, $C_2$ is the Gompertz asymptote level, and $\tau$ is a parameter denoting the start of infectiousness, assumed to be one day after infection within the incubation period. For each infected individual, the incubation period was sampled independently from a LogNormal distribution, with parameters 1.434 (shape) and 0.661 (rate), having a mean of 5.2 days. Next, for each value of $\tau$, the parameters $n, C_1,$ and $C_2$ were estimated using a least-squares optimization procedure with the objective function:

$$\min_{n,\,C_1,\,C_2} \sum_{t=1}^{26} (\rho(t) - [s_\tau(t) g(t-\tau)])^2$$

where $\rho(t)$ is the percent positivity data of 209 COVID-19 patients for 26 days after the start of symptoms, including the day of symptom onset (Figure A2, brown circles). The fitted sensitivity function determined the probability of being detected at the time of a nasopharyngeal test. An example of this fitting process when $\tau = 4.2$ (corresponding to an average incubation period of 5.2 days minus 1 day to the start of infectiousness) is shown in Figure A2 (red curve).

**Normalization process for determining temporal sensitivity of saliva testing**

To determine the sensitivity curve for the saliva test, we used recent empirical studies for the estimates of saliva testing sensitivity in the range of 70% - 97% [3-5]. Since viral load in saliva samples have shown to be comparable to NP samples over time [6-8], we assumed that the sensitivity curve of saliva testing has a similar shape to the sensitivity curve of NP testing. We therefore applied this sensitivity range of saliva test to the normalized sensitivity curves of NP testing. The normalized sensitivity curve of NP testing was calculated by

$$s_\tau^{\text{norm}}(t) = \frac{s_\tau(t) - \min_t(s_\tau(t))}{\max_t(s_\tau(t)) - \min_t(s_\tau(t))}$$

Note that $\min_t(s_\tau(t)) = 0$. Next, each normalized curve was multiplied by a number sampled from a uniform distribution with parameters $a = 0.70$ and $b = 0.97$, corresponding to the estimated sensitivity range for saliva testing. The mean and the range for these curves are shown in Figure A2 (black curve and shaded area) when the incubation period was at the average of 5.2 days.

**Calculation of the average probability of detection**

The probability of identifying an infected individual was calculated by

$$P = 1 - (1 - p_1)(1 - p_2) \cdots (1 - p_k)$$

where $p_i, i = 1, \cdots, k$ is the probability of being detected by a test administered at time $t_i$, and $k$ is the total number of tests administered during the infectious period or the pre-symptomatic stage. The probability $p_i$ was calculated by evaluating the sensitivity function at time $t_i$ for both NP and saliva testing. The case detection percentage was then estimated as the average of probabilities $P$ for all individuals in each simulation. We generated the mean detection distributions by running 500 Monte-Carlo simulations.

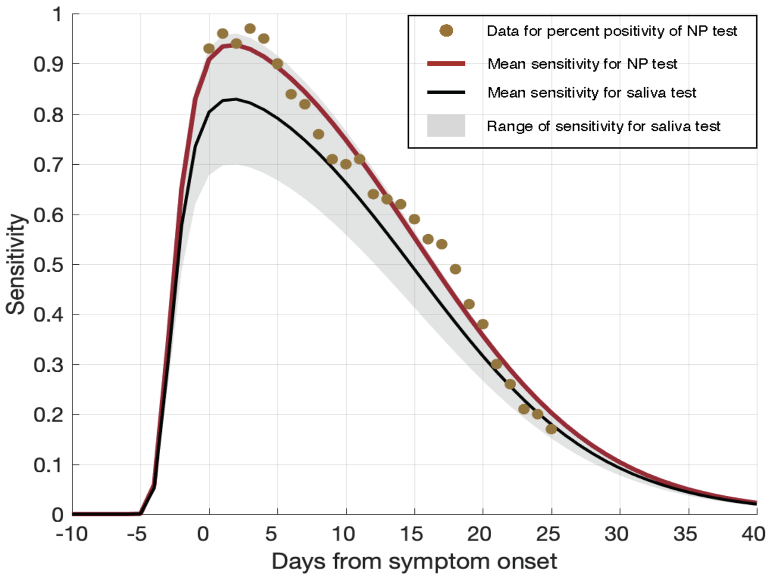


**Figure A2.** Fitting of the modified Hill function, based on the infectiousness profile, to the percent positivity data of the nasopharyngeal RT-PCR test for 209 COVID-19 patients [9] with an average incubation period of 5.2 days [10]. The red curve shows the fit to data for the percent positivity of nasopharyngeal test (brown circles) post-symptom onset, and the shaded area represents the sensitivity of a saliva test, with the mean illustrated by the black curve.

### References


1. He X, Lau EHY, Wu P, et al. Temporal dynamics in viral shedding and transmissibility of COVID-19. *Nature Medicine*. 2020;26(5):672-675.
2. Ashcroft P, Huisman JS, Lehtinen S, et al. COVID-19 infectivity profile correction. *Swiss Medical Weekly*. 2020;150(3132).
3. Yokota I, Shane PY, Okada K, et al. Mass screening of asymptomatic persons for SARS-CoV-2 using saliva. *Clin Infect Dis*. Published online September 25, 2020.
4. Jamal AJ, Mozafarihashjin M, Coomes E, et al. Sensitivity of Nasopharyngeal Swabs and Saliva for the Detection of Severe Acute Respiratory Syndrome Coronavirus 2. *Clin Infect Dis*. Published online June 25, 2020.
5. Wyllie AL, Fournier J, Casanovas-Massana A, et al. Saliva or Nasopharyngeal Swab Specimens for Detection of SARS-CoV-2. *N Engl J Med*. 2020;383(13):1283-1286.
6. Larremore DB, Wilder B, Lester E, et al. *Test Sensitivity Is Secondary to Frequency and Turnaround Time for COVID-19 Surveillance*. medRxiv; 2020.
7. Iwasaki S, Fujisawa S, Nakakubo S, et al. Comparison of SARS-CoV-2 detection in nasopharyngeal swab and saliva. *J Infect*. 2020;81(2):e145-e147.
8. Kim SE, Lee JY, Lee A, et al. Viral Load Kinetics of SARS-CoV-2 Infection in Saliva in Korean Patients: a Prospective Multi-center Comparative Study. *J Korean Med Sci*. 2020;35(31):e287.
9. Miller TE, Beltran WFG, Bard AZ, et al. Clinical sensitivity and interpretation of PCR and serological COVID-19 diagnostics for patients presenting to the hospital. *The FASEB Journal*. 2020;34(10):13877-13884.

10. Li Q, Guan X, Wu P, et al. Early Transmission Dynamics in Wuhan, China, of Novel Coronavirus–Infected Pneumonia. *N Engl J Med*. 2020;382(13):1199-1207.
